# It Takes Two to Tango: Combining Conventional Culture with Molecular Diagnostics Enhances Accuracy of *Streptococcus pneumoniae* Detection and Pneumococcal Serogroup/Serotype determination in Carriage

**DOI:** 10.1101/2021.11.26.21266900

**Authors:** Willem R. Miellet, Janieke van Veldhuizen, David Litt, Rob Mariman, Alienke J. Wijmenga-Monsuur, Paul Badoux, Tessa Nieuwenhuijsen, Rebecca Thombre, Sanaa Mayet, Seyi Eletu, Carmen Sheppard, Marlies A. van Houten, Nynke Y. Rots, Elizabeth Miller, Norman K. Fry, Elisabeth A.M. Sanders, Krzysztof Trzciński

## Abstract

**Background:** The specificity of molecular methods for the detection of *Streptococcus pneumoniae* carriage is under debate. We propose a procedure that increases the accuracy of molecular detection of live pneumococci in polymicrobial respiratory samples.

**Methods:** Culture and qPCR methods were applied to detect *S. pneumoniae* and pneumococcal serotypes in 1549 nasopharyngeal samples collected in the Netherlands (n=972) and England (n=577) from 946 toddlers and 603 adults, and in paired oropharyngeal samples collected exclusively from 319 Dutch adults. Samples with no live pneumococci isolated at primary diagnostic culture yet generating pneumococcus-specific signal in qPCRs were re-examined with a second, qPCR-guided culture. Optimal C_q_ cut-offs for positivity in qPCRs were determined via receiver operating characteristic (ROC) curve analysis using isolation of live pneumococci from the primary and qPCR-guided cultures as reference.

**Results:** Detection of *S. pneumoniae* and pneumococcal serotypes with qPCRs in cultured (culture-enriched) nasopharyngeal samples exhibited near-perfect agreement with conventional culture (Cohen’s kappa: 0.95). Molecular methods also displayed increased sensitivity of detection for multiple serotype carriage. Among paired samples from adults, the sensitivity of *S. pneumoniae* detection in primary nasopharyngeal plus oropharyngeal cultures was significantly lower compared with molecular detection in both culture-enriched samples together (*p*<0.0001) and also in culture-enriched oropharyngeal samples alone (*p*<0.05).

**Conclusions:** The sensitivity of *S. pneumoniae* carriage surveillance can be greatly improved by complementing conventional culture with qPCR and *vice versa*. The specificity of molecular methods for the detection of live pneumococci can be enhanced by incorporating statistical procedures based on ROC curve analysis. The procedure we propose improves detection of *S. pneumoniae* carriage in adults in particular and enhances specificity of serotype carriage detection.

## INTRODUCTION

*Streptococcus pneumoniae* (pneumococcus) is the most common aetiological agent of invasive bacterial disease [1] and of community-acquired pneumonia of bacterial aetiology [2]. Despite being vaccine-preventable, pneumococcal disease remains among the leading causes of death in childhood [3] as available vaccines target only a various subsets of only twenty-four from ca. 100 of the known serotypes. Due to high pneumococcal carriage rates [4, 5] children are considered the primary reservoir of pneumococcus and the main drivers of transmission and infections in any population [6–8]. Children are also the primary group targeted with pneumococcal vaccination [9].

With pneumococcal vaccines protecting not only against disease but also against colonization, carriage is now an accepted endpoint in vaccination studies [10]. Following the introduction of pneumococcal conjugate vaccines (PCV), of which ten-valent (PHiD-CV, GSK) and thirteen-valent (PCV13, Pfizer) are currently marketed, epidemiological surveillance of carriage became an essential tool for monitoring the direct and indirect (herd protection) effects of vaccination. Surveillance of carriage is also used to detect the emergence and track expansion of non-vaccine serotypes, the phenomenon described as vaccine-induced serotype replacement [11]. Finally, carriage studies are instrumental in monitoring serotype-associated invasiveness [12].

To understand the trends in pneumococcal epidemiology and guide strategies for PCV use, it is critical to establish methodology for *S. pneumoniae* carriage detection that is sufficiently sensitive across all ages [13, 14]. However, conventional culture, which is considered to be the gold standard method in carriage detection, is not suitable for detection of multiple serotype carriage [15] and lacks the sensitivity when used to detect *S. pneumoniae* in age groups other than children [16]. There is also evidence that testing solely a single site within the upper airways reduces sensitivity of carriage detection [17–19].

Molecular methods have largely improved the sensitivity of *S. pneumoniae* detection and have made multiple serotype carriage detection feasible [4, 20]. Using assays developed by our groups and others, we have tested the serotype composition of respiratory samples from children and adults and have demonstrated an under-detection of *S. pneumoniae* and of individual pneumococcal serotypes by culture when compared with molecular methods [7, 18, 21–24]. However, some caution that molecular methods exhibit poor specificity due to the presence of pneumococcal genes among commensal streptococci [25–27]. It could also be argued that molecular detection is unable to discriminate between live bacteria and presence of relic DNA [28].

Here, we outline the protocol that combines conventional culturing with pneumococcus-specific qPCRs and employs statistical procedures to interpret the molecular results and enhances the specificity of the molecular methods. We show that molecular methods applied to nasopharyngeal samples demonstrate near-perfect agreement with primary culture and yet the sensitivity of *S. pneumoniae* carriage surveillance can be greatly enhanced by complementing conventional culture with qPCRs and *vice versa*. We also show that testing nasopharyngeal samples alone leads to underestimation of pneumococcal carriage in adults.

## MATERIALS AND METHODS

### Study Design and Ethics Statement

Pneumococcal carriage was investigated in two cross-sectional prospective observational studies conducted in 2015/2016 in community-dwelling individuals in the Netherlands [29] and in England [30]. The study conducted in the Netherlands was approved by the Medical Ethics Committee Noord Holland (NTR5405 on http://www.trialregister.nl). The study conducted in England was approved by the NHS Health Research Authority and the London Fulham Research Ethics Committee (reference 15/LO/0458) and was registered on clinicaltrials.gov (reference NCT02522546). Both studies were conducted in accordance with Good Clinical Practice and the Declaration of Helsinki.

### Sample Collection and Laboratory Processing

Respiratory samples were collected in the Netherlands between October 2015 and March 2016 in the study on carriage of respiratory bacterial pathogens coordinated by National Institute of Public Health and the Environment. Nasopharyngeal samples were collected in children aged 24 months (±1 month) vaccinated with PHiD-CV according to “2 primary + 1 booster” (2p+1b) dose schedule, children aged 44 to 49 months vaccinated with PHiD-CV in 3p+1b schedule, and parents of the 24-month-old children (one parent per child). Oropharyngeal swabs were also collected from Dutch adults [29, 31]. The English study took place between July 2015 and June 2016 and was conducted by the National Vaccine Evaluation Consortium which included Public Health England [30]. In England, nasopharyngeal samples were collected in children aged 1-5 years vaccinated with PCV13 according to a 2p+1b schedule and in their household contacts (adults aged >20 years).

All samples were obtained according to World Health Organization standard procedures [4]. Immediately after sampling, swabs in the Netherlands were placed in liquid Amies transport medium (Copan, Brescia, Italy) and within 8 hours transported to the diagnostic laboratory. In England swabs were placed in skim-milk, tryptone, glucose and glycerol (STGG) broth and delivered to a diagnostic laboratory within 48 hours (**Table S1**).

### Conventional Culture

Approximately 50 µl of specimen was used as an inoculum in detection of pneumococci by conventional culture (primary culture). In the Netherlands samples were cultured on SB7-Gent agar selective for streptococci (BA-GENT, Oxoid, Badhoeve Dorp, The Netherlands) and on non-selective Columbia blood agar (CBA, Oxoid). In England samples were cultured on Streptococcus-selective Blood Agar (COBA, Oxoid, Basinstoke, United Kingdom) and CBA (Oxoid). Remaining of transport media were stored frozen at −70°C. In the Netherlands, samples were aliquoted into 200 μl volumes, and a single aliquot was supplemented with glycerol (10% v/v final concentration) prior to freezing.

After overnight incubation at 37°C and 5% CO_2_, cultures were screened for pneumococcus-like colonies to be re-cultured. Once screened, in the Netherlands all colony growth was harvested from BA-GENT plate into 10% glycerol in brain heart infusion broth (Oxoid) [24]. In England, all colony growth was harvested from any plates containing alpha-haemolytic colonies into PBS, centrifuged, the supernatant removed, and any growth stored as a pellet [30]. These samples were considered culture-enriched (CE) for pneumococci and stored at −80°C. Cultured strains (one per sample, but more if distinct pneumococcal morphotypes were apparent) were serotyped by Quellung method in the Netherlands [29] and with the previously described PneumoCaT bioinformatic pipeline in England that was supplemented with slide agglutination when required [30, 32].

### Molecular Detection of Streptococcus pneumoniae

Molecular detection of *S. pneumoniae* in respiratory samples was conducted as described previously [24]. For this, Amies medium, STGG medium, and culture-enriched samples were shipped on dry ice from primary diagnostic sites to the study central laboratory in the Netherlands. There, pellets of culture-enriched samples collected in England were reconstituted to the original volume. Next, nucleic acids were extracted from 100 µl of medium using the DNeasy Blood and Tissue Kit (Qiagen) and eluted into 200 µl. These extractions represented minimally processed samples. For culture-enriched samples, 100 µl of a bacterial growth harvest was centrifuged for 2 min at 14,000 ×g, the pellet was resuspended with 90 µl of TE buffer (20 mM Tris-HCl [pH 8.0], 2 mM EDTA) and incubated for 15 min at 95°C. Next, 90 µl of lysis buffer (20 mM Tris-HCl [pH 8.0], 2 mM EDTA, 2.4% Triton X-100 and 40 mg/ml lysozyme) was added, and the samples were processed as above. Pneumococcal DNA was detected via single plex qPCR using primers and probe targeting sequences within genes coding for the pneumococcal iron uptake ABC transporter lipoprotein PiaB [18], and for the major pneumococcal autolysin LytA [33] by testing 5.5 μl DNA from minimally processed or 1.0 μl of culture-enriched samples.

### Molecular Detection of Pneumococcal Serotypes

DNA extracted from culture-enriched harvests was used to determine the serotype composition of nasopharyngeal and oropharyngeal samples using a panel of primers and probes [34–36], targeting all serotypes covered by pneumococcal vaccines available either in the Netherlands or in England at the time of sample collection (PHiD-CV, PCV13 and 23-valent polysaccharide vaccine, PPV23, Merc Sharp Dohme) except for serotype 2. The panel also covered selected non-vaccine serotypes (serotypes 6C, 6D, 7A, 10B, 12A, 12B, 15A, 15C, 15F, 16F, 21, 22A, 23A, 23B, 33A, 34, 35B, 35F, 37 and 38). However, with certain assays targeting more than one serotype it was not possible to distinguish between serotypes 6A and 6B; 6C and 6D; 7A and 7F; 9A, 9L, 9N and 9V; 10A and 10B; 11A and 11D; 12A, 12B and 12F; 15A, 15B, 15C and 15F; 33A, 33F and 37; 35B and 35C when detected with qPCR. A sample pooling strategy was employed when testing for serotypes in order to reduce the number of serotype-specific qPCRs. For this, samples generating any signal below C_q_ 40 for either *piaB* or *lytA* were pooled in groups of five to be tested [7] and all remaining samples were pooled in groups of ten. Samples negative for *piaB* and *lytA* were tested to assess specificity of serotype/serogroup-specific assays [7]. Samples from pools generating a signal for a particular serotype/serogroup were tested individually. In the Netherlands, the PCRs were performed on the LightCycler 2 platform and in England they were performed on the QuantStudio 7 platform, using identical PCR conditions.

### qPCR-guided Recovery of Live *S. pneumoniae* Strains

Culture-enriched samples classified as negative in primary diagnostic culture but positive by qPCR were revisited with culture in a second attempt to isolate live pneumococci. For this, CBA plates were inoculated with 100 µl of 10^-1^-10^-3^ and 10^-3^-10^-5^ dilution of culture-enriched nasopharyngeal and culture-enriched oropharyngeal samples, respectively, and incubated at 37°C and 5% CO_2_. Pneumococcus-like colonies were individually tested in qPCR for *piaB* and *lytA* and confirmed to be *S. pneumoniae* based on susceptibility to optochin.

## Statistical Analysis

Data analysis was performed in GraphPad Prism software version 9.1.0 and R version 4.1.0. Receiver operating characteristic (ROC) curve analysis was performed using the “cutpointr” R package to validate qPCR results with culture results (primary culture plus qPCR-guided culture). Maximum Youden index values, the sum of sensitivity and specificity minus one, were estimated via bootstrapping (n=1000) on *piaB* and *lytA* qPCR data from respiratory samples to determine optimal cut-off values for qPCR detection [37]. Two-way mixed effects intraclass correlations (ICCs) [38] and Bland-Altman plots [39] were used to evaluate agreement for quantitative results using the “ïrr” and “blandr” R packages, respectively. Carriage rates were compared with McNemar’s test unless otherwise stated. Estimates for accuracy of diagnostic tests between methods (subgroups) were compared with a test of interaction [40]. A *p* value of <0.05 was considered significant. Diagnostic test parameters (predictive values, sensitivity, and specificity) were calculated using an in-house made R function (https://github.com/wmiellet/test-comparison-R), and 95% confidence intervals were calculated with Wilson-Brown score. Cohen’s kappa, a measure of chance-corrected agreement, and its 95% confidence interval was calculated as described by McHugh [41] and ratios of ≤ 0, 0.01-0.20, 0.21-0.40, 0.41-0.60, 0.61-0.80 and >0.81 were interpreted as exhibiting poor, slight, fair, moderate, substantial, and near-perfect agreement, respectively [42]. For comparison between serotyping by culture and by qPCR, analysis was limited to qPCR-targeted serotypes. For serogroup-specific qPCR assays a result was considered congruent when a serogroup was detected in qPCR that matched the serogroup of the serotype detected by culture.

### Assessment of method’s inter-laboratory reproducibility

One-hundred seventy-six nasopharyngeal samples collected in England and n=59 oropharyngeal samples collected in the Netherlands have been selected to evaluate the inter-laboratory reproducibility of molecular methods. For this, an aliquot of minimally processed STGG nasopharyngeal sample, a fresh preparation of culture enriched nasopharyngeal sample from a subset of the English study samples and culture-enriched oropharyngeal samples from a subset of the Dutch study samples were tested at the study site in England, as described above. Results for paired samples were compared between centres by calculating the percent agreement and Cohen’s kappa. Quantitative results of both laboratories were also compared by calculating an intraclass correlation coefficient (ICC) and by comparing results in Bland-Altman plots. Carriage rates between both laboratories were compared using Cohen’s kappa.

## RESULTS

We evaluated the performance of conventional and molecular methods applied to detect *S. pneumoniae* and pneumococcal serotypes in 1549 nasopharyngeal samples collected from 946 children aged 1-5 years (n=653 in the Netherlands and n=293 in England) and from 603 adults (n=319 in the Netherlands and n=284 in England), and on oropharyngeal samples from 319 adults in the Netherlands. All PCR results shown were generated by testing the Dutch and English samples in the Dutch laboratory, except were stated otherwise.

### Detection of Streptococcus pneumoniae

When analysing culturing results without the addition of PCR-guided culturing, live *S. pneumoniae* was isolated from 29% (445/1549) of primary cultures of nasopharyngeal swabs (**Fig. 1A**), with significantly higher fraction of samples from children (45% or 422/946) compared with adults (4% or 23/603) positive for pneumococcus (Pearson’s chi-square, *p*<0.0001). Among 319 adults sampled in the Netherlands (**Fig. 1B**), live pneumococcus was significantly more often cultured from nasopharyngeal (n=15 or 5%) compared with oropharyngeal (n=3 or 1%) samples (*p*<0.01).

**Figure 1.**
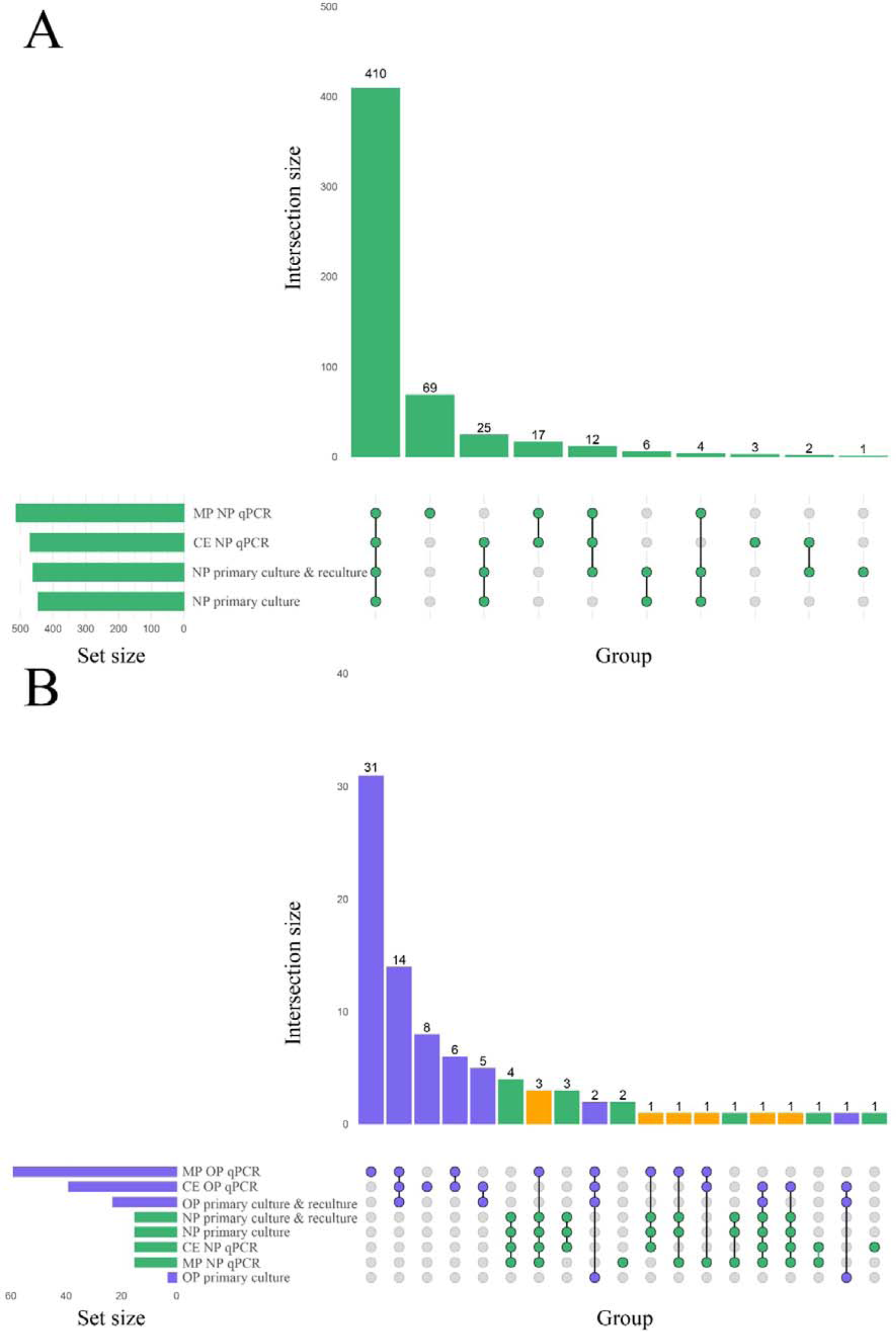
(**A**) Matrix layout for observed intersections of the *Streptococcus pneumoniae* detection procedures applied to n=1549 nasopharyngeal samples, sorted by size. (**B**) Matrix layout for observed intersections of the *S. pneumoniae* detection procedures applied to nasopharyngeal and oropharyngeal samples from n=319 adults sampled in the Netherlands, sorted by size. Circles in the matrix indicate sets that are part of the intersection, and circles are coloured by sample type with nasopharyngeal and oropharyngeal samples coloured in green and blue, respectively. The bar diagram displaying the intersection size is coloured using the same scheme, with orange indicating an intersection that represents positivity in both sample types.

Next, we conducted qPCRs on minimally processed and culture-enriched samples. To enhance the specificity of detection we used a “two-to-tango” approach by quantifying *piaB* and *lytA* genes and considering a sample to be positive for *S. pneumoniae* when both targets were detected. When applying an arbitrary quantification cycle (^A^C_q_) of <40 C_q_ as criterium for positivity altogether 583 nasopharyngeal swabs (38% of 1549) were identified as positive for pneumococcus (**Fig. 2A** and **2B**). The fraction of positive samples was significantly higher among minimally processed compared with culture-enriched swabs (561 or 36% versus 479 or 31% of 1549; *p*<0.0001). In line with results of primary diagnostic culture, here too the proportion of positive samples was higher in children compared with adults (Pearson’s chi-square, *p*<0.0001) whenever nasopharyngeal swabs were tested minimally processed (56% or 528/946 versus 5% or 33/603) or culture-enriched (48% or 454/946 versus 4% or 25/603). However, contrary to results of primary diagnostic culture, among adults significantly larger proportions of oropharyngeal compared with nasopharyngeal samples (*p*<0.0001) have been classified as positive for pneumococcus with qPCR whenever tested minimally processed (26% or 83/319 versus 6% or 19/319) or culture-enriched (18% or 58/319 versus 6% or 18/319).

**Figure 2.**
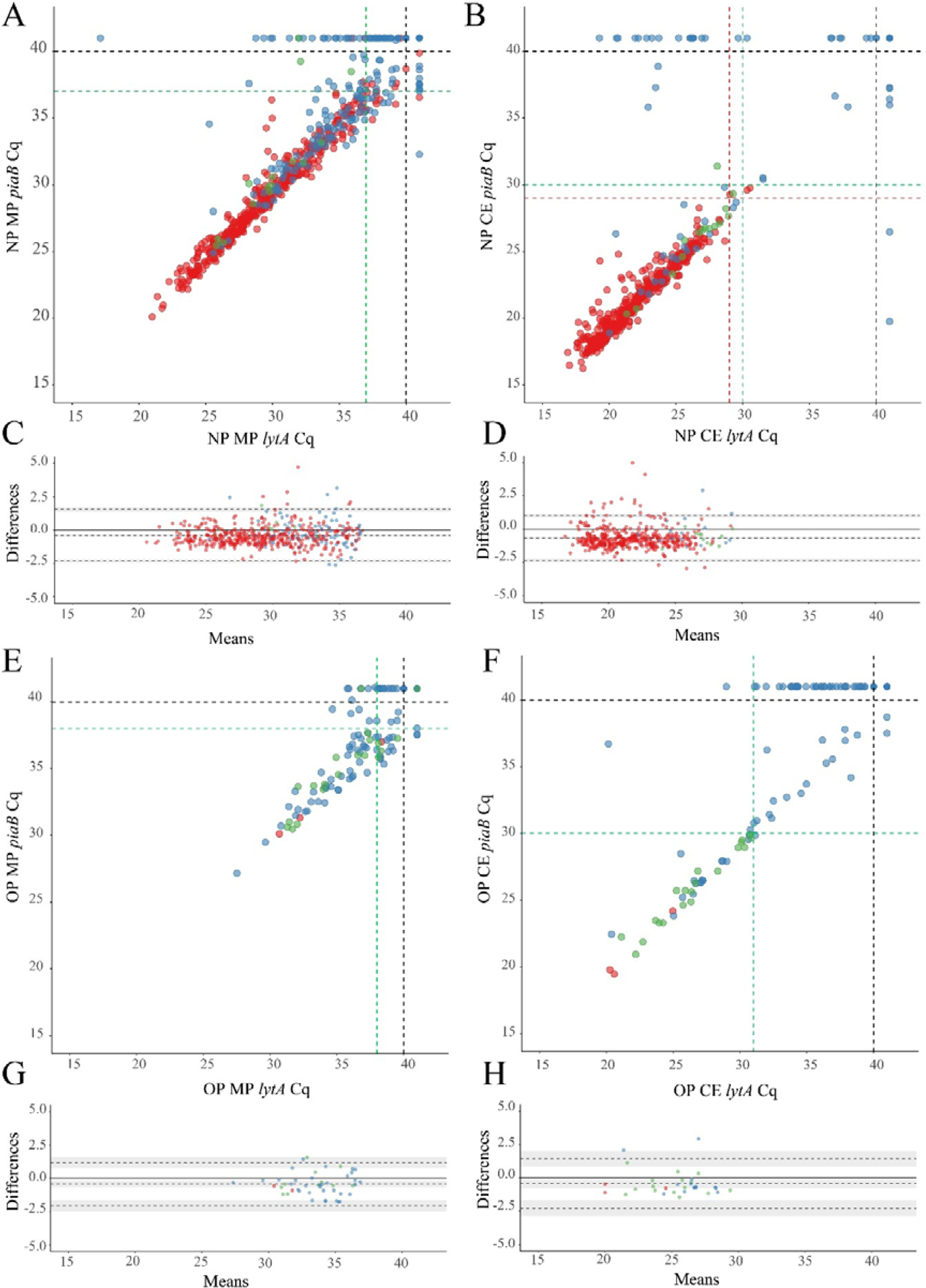
Scatter plots (**A**, **B**, **E** and **F**) displaying correlation between cycle threshold (C_q_) from real-time PCR assays targeting the *Streptococcus pneumoniae* genes *piaB* and *lytA* for minimally processed (**A** and **E**) and culture-enriched (**B** and **F**) nasopharyngeal (**A** and **B**) and oropharyngeal (**E** and **F**) samples followed by the Bland-Altman plots (**C**, **D**, **G** and **H**) displaying agreement between *piaB* and *lytA* for samples positive with minimally processed (**C** and **G**) and culture-enriched (**D** and **H**) ^ROCd^C_q_ criteria in nasopharyngeal (**C** and **D**) and oropharyngeal (**G** and **H**) samples. The shaded grey areas mark the 95% confidence interval of the upper limit of agreement, bias and lower limit of agreement. The Bland-Altman plots show the absence of systematic bias between *piaB* and *lytA* because the line of equality (grey solid line) is within the limits of agreement. Bias between *piaB* and *lytA* is more often observed in high C_q_ samples. Primary culture-positive and qPCR-guided culture-positive samples are indicated with red and green dots, respectively. Culture-negative samples are indicated with blue dots. In panels **A**, **B**, **E** and **F** black dashed lines indicate the arbitrary C_q_ criterium (<40 C_q_), green dashed lines indicate ^ROCd^C_q_ criteria derived by validating qPCR results with primary and qPCR-guided cultures results, and red dashed lines in panels **A** and **B** indicate ^ROCd^C_q_ criteria derived by validating qPCR results with primary culture alone.

Next, samples negative for pneumococcus in primary diagnostic cultures yet generating signal in qPCRs for *piaB* and *lytA* were cultured again. This qPCR-guided culture effort increased the number of samples from which live pneumococci was cultured, in relatively small albeit significant increase of 3% (from 445 vs. 460 of 1549; *p<*0.0001) for nasopharyngeal (**Fig. 1A, 2A**, and **2B**) and 666% increase (from 3 to 23 of 319; *p*<0.0001) for oropharyngeal samples (**Fig. 1B, 2E**, and **2F**).

To validate qPCR results using culture and to further enhance the specificity of qPCR methods for the detection of live pneumococci we performed receiver operating characteristic (ROC) curve analysis and identified C_q_ cut-off values that yielded maximum Youden indices (**Table 1**). Reliability of positive qPCR results was assessed with Bland-Altman plots (**Fig. 2C** and **2D** and **Fig. 2G** and **2H** and supplementary information **Table S2**) and by calculating the intraclass correlation coefficient between *piaB* and *lytA*. Samples with C_q_ values below the ROC-derived C_q_ criterium (^ROCd^C_q_) demonstrated excellent agreement between *piaB* and *lytA* while samples with a C_q_ value above the ^ROCd^C_q_ threshold displayed poor agreement (**Table S2**). Using the ^ROCd^C_q_ criterium we classified 33% (512/1549) of minimally processed nasopharyngeal samples as positive for pneumococcus (**Fig. 1A**). It included 52% (489/946) samples from children and 4% (23/603) from adults. For culture-enriched samples application of ^ROCd^C_q_ resulted in 30% (469/1549) classified as positive including 47% (447/946) samples from children and 4% (22/603) from adults (**Fig. 1A**). With ^ROCd^C_q_, the adults in the Netherlands were again significantly more often positive for pneumococcus in oropharyngeal compared to nasopharyngeal samples whenever tested minimally processed (19% or 59/319 versus 5% or 15/319; *p*<0.0001) or culture-enriched (12% or 39/319 versus 5% or 15/319; *p*<0.01) (**Fig. 1B**).

**Table 1.** Optimal qPCR cycle threshold C_q_ and corresponding parameters for *Streptococcus pneumoniae* carriage detection in nasopharyngeal (n=1549) and oropharyngeal (n=319) samples. Results from qPCR were validated in a receiver operating characteristic curve analysis with culture as reference.

| Parameter | Sample | Reference | Optimal threshold<br>*piaB*<br>(95% CI) | Youden index<br>*piaB* | Sensitivity<br>*piaB* | Specificity<br>*piaB* | Optimal threshold<br>*lytA*<br>(95% CI) | Youden index<br>*lytA* | Sensitivity<br>*lytA* | Specificity<br>*lytA* |
| --- | --- | --- | --- | --- | --- | --- | --- | --- | --- | --- |
| Minimally processed | NP | Primary cultures | 37.13<br>(36.49 – 37.76) | 0.85 | 0.95 | 0.90 | 37.47<br>(36.44 – 38.39) | 0.82 | 0.94 | 0.87 |
|  | NP | Primary culture & reculture | 37.21<br>(36.51 – 38.46) | 0.85 | 0.95 | 0.90 | 37.29<br>(36.52 – 38.61) | 0.83 | 0.94 | 0.89 |
|  | OP | Primary culture & reculture | 38.73<br>(37.29 – 39.39) | 0.69 | 0.91 | 0.78 | 37.47<br>(36.52 – 37.66) | 0.69 | 0.87 | 0.82 |
| Culture-enriched | NP | Primary cultures | 28.71<br>(26.59 – 29.69) | 0.94 | 0.98 | 0.97 | 29.19<br>(27.41 – 30.25) | 0.93 | 0.98 | 0.95 |
|  | NP | Primary culture & reculture | 30.26<br>(29.91 – 30.47) | 0.96 | 0.98 | 0.98 | 29.99<br>(28.91 – 30.47) | 0.94 | 0.98 | 0.96 |
|  | OP | Primary culture & reculture | 29.78<br>(28.84 – 29.93) | 0.91 | 0.96 | 0.95 | 30.73<br>(29.42 – 30.81) | 0.85 | 0.91 | 0.94 |

With *S. pneumoniae* isolated from only three samples there was insufficient statistical power to perform the receiver operating characteristic curve analysis on oropharyngeal samples with primary cultures as a reference.

### Comparison of Molecular Methods to Culturing Live *S. pneumoniae*

We compared the diagnostic accuracy of detection methods using the combined results of primary diagnostic and qPCR-guided culturing as reference and applying ^ROCd^C_q_ criteria for positivity in qPCRs (**Fig. 1A**). For nasopharyngeal samples alone (**Table 2**), molecular detection of pneumococcus displayed near-perfect, and substantial agreement to the reference for culture-enriched and minimally processed samples, respectively. This represented significantly reduced agreement for minimally processed compared with culture-enriched samples *(p*<0.0001). Molecular detection in minimally processed samples identified significantly more samples positive for pneumococcus compared with culture-enriched samples from children (*p*<0.0001) but not adults (*p*=1), the difference we attributed to low number of positive nasopharyngeal samples among collected from adults. Exclusively for children, molecular detection of pneumococcus in culture-enriched samples demonstrated significantly increased sensitivity (*p*<0.001) and specificity (*p*<0.0001) compared with molecular detection in minimally processed samples.

**Table 2.** The accuracy of *Streptococcus pneumoniae* detection in n=1549 nasopharyngeal samples from children (n=946) and adults (n=603) tested using conventional diagnostic culture and molecular methods applied to DNA extracted from minimally processed and culture-enriched samples and applying ^ROCd^C_q_ thresholds for a sample positivity in qPCRs. Measures of diagnostic accuracy were calculated by comparing the number of detected samples positive per method with the number of (n=460 for all individuals, and n=437 and n=23 for nasopharyngeal samples from children and adults, respectively) individuals positive for *S. pneumoniae* based on isolation of live pneumococcus either from the primary diagnostic or qPCR-guided culture.

| Method | Study population | Percent (n) of positive samples (95%CI) | PPV % (95%CI) | NPV % (95%CI) | Sensitivity % (95%CI) | Specificity % (95%CI) | Concordance % (95%CI) | $\kappa$ (95%CI) |
| --- | --- | --- | --- | --- | --- | --- | --- | --- |
| primary culture | All | 28.7 (445)<br>(26.5 – 31.0) | 100<br>(99.1 – 100) | 98.6<br>(97.8 – 99.2) | 96.7<br>(94.7 – 98.0) | 100<br>(99.6 – 100) | 99.0<br>(98.4 – 99.4) | 0.98<br>(0.96 – 0.99) |
|  | Children | 44.6 (422)<br>(41.5 – 47.8) | 100<br>(99.1 – 100) | 97.1<br>(95.3 – 98.3) | 96.6<br>(94.4 – 97.9) | 100<br>(99.3 – 100) | 98.4<br>(97.4 – 99.0) | 0.97<br>(0.95 – 0.98) |
|  | Adults | 3.8 (23)<br>(2.6 – 5.7) | 100<br>(85.7 – 100) | 100<br>(99.3 – 100) | 100<br>(85.7 – 100) | 100<br>(99.3 – 100) | 100<br>(99.4 – 100) | 1 |
| qPCRs on minimally processed samples | All | 33.1 (512)<br>(30.8 – 35.4) | 83.2<br>(79.7 – 86.2) | 96.7<br>(95.5 – 97.6) | 92.6<br>(89.8 – 94.7) | 92.1<br>(90.3 – 93.6) | 92.3<br>(90.8 – 93.5) | 0.82<br>(0.79 – 0.85) |
|  | Children | 51.7 (489)<br>(48.5 – 54.9) | 83.6<br>(80.1 – 86.7) | 93.9<br>(91.3 – 95.7) | 93.6<br>(90.9 – 95.5) | 84.3<br>(80.9 – 87.2) | 88.6<br>(86.4 – 90.5) | 0.77<br>(0.73 – 0.81) |
|  | Adults | 3.8 (23)<br>(2.6 – 5.7) | 73.9<br>(53.5 – 87.5) | 99.0<br>(97.8 – 99.5) | 73.9<br>(53.5 – 87.5) | 99.0<br>(97.8 – 99.5) | 98.0<br>(96.6 – 98.9) | 0.73<br>(0.58 – 0.88) |
| qPCRs on culture-enriched samples | All | 30.3 (469)<br>(28.0 – 32.6) | 95.7<br>(93.5 – 97.2) | 99.0<br>(98.2 – 99.4) | 97.6<br>(95.8 – 98.7) | 98.2<br>(97.2 – 98.8) | 98.0<br>(97.2 – 98.6) | 0.95<br>(0.94 – 0.97) |
|  | Children | 47.3 (447)<br>(44.1 – 50.4) | 96.0<br>(93.7 – 97.4) | 98.4<br>(96.9 – 99.2) | 98.2<br>(96.4 – 99.1) | 96.5<br>(94.5 – 97.8) | 97.3<br>(96.0 – 98.1) | 0.94<br>(0.92 – 0.97) |
|  | Adults | 3.7 (22)<br>(2.4 – 5.5) | 90.9<br>(72.2 – 97.5) | 99.5<br>(98.5 – 99.8) | 87.0<br>(67.9 – 95.5) | 99.7<br>(98.8 – 99.9) | 99.2<br>(98.1 – 99.6) | 0.88<br>(0.78 – 0.99) |

PPV – positive predictive value; NPV – negative predictive value; 95%CI – 95% confidence interval; $\kappa$ – Cohen's kappa where $\leq 0$ , 0.01-0.20, 0.21-0.40, 0.41-0.60, 0.61-0.80, $>0.81$ are interpreted as poor agreement, slight, fair, moderate, substantial, and almost perfect agreement, respectively.

For paired nasopharyngeal and oropharyngeal samples from adults, the isolation of *S. pneumoniae* in primary or qPCR-guided culture in either nasopharyngeal or oropharyngeal swab was used as the reference in method accuracy analysis (**Table 3**). Although the sensitivity of *S. pneumoniae* detection with primary nasopharyngeal culture was significantly higher compared with corresponding values for primary oropharyngeal cultures (*p*<0.001), every detection method when applied to a single sample type displayed only slight to moderate agreement to the reference. Moreover, the sensitivity of *S. pneumoniae* detection with either nasopharyngeal and oropharyngeal primary cultures was significantly lower compared with molecular detection in culture-enriched oropharyngeal samples (*p*<0.05) and also when compared with molecular detection with either culture-enriched sample of either type (*p*<0.0001). To this extent, molecular detection in culture-enriched oropharyngeal plus nasopharyngeal samples displayed the highest agreement to the reference out of all other evaluated approaches (**Table 3**).

**Table 3.** The accuracy of *Streptococcus pneumoniae* detection in paired nasopharyngeal and oropharyngeal samples from n=319 adults tested using molecular methods applied to DNA extracted from minimally processed and culture-enriched samples and applying ^ROCd^C_q_ thresholds for a sample positivity in qPCRs. Measures of diagnostic accuracy were calculated by comparing the number of detected samples positive per method with the overall number of n=37 individuals identified as carriers of *S. pneumoniae* based on isolation of live pneumococcus either at the primary diagnostic or qPCR-guided culture and either from nasopharyngeal or oropharyngeal sample.

| Method | Positivity in | Percent (n) of positive samples<br>(95%CI) | PPV<br>%<br>(95%CI) | NPV<br>%<br>(95%CI) | Sensitivity<br>%<br>(95%CI) | Specificity<br>%<br>(95%CI) | Concordance<br>%<br>(95%CI) | $\kappa$<br>(95%CI) |
| --- | --- | --- | --- | --- | --- | --- | --- | --- |
| Primary culture alone | NP | 4.7 (15)<br>(2.9 – 7.6) | 100<br>(79.6 – 100) | 92.8<br>(89.3 – 95.2) | 40.5<br>(26.3 – 56.5) | 100<br>(98.7 – 100) | 93.1<br>(89.8 – 95.4) | 0.55<br>(0.36 – 0.73) |
|  | OP | 0.9 (3)<br>(0.3 – 0.7) | 100<br>(43.9 – 100) | 89.2<br>(85.3 – 92.2) | 8.1<br>(2.8 – 21.3) | 100<br>(98.7 – 100) | 89.3<br>(85.5 – 92.3) | 0.13<br>(-0.14 – 0.41) |
|  | either NP or OP | 5.6 (18)<br>(3.6 – 8.7) | 100<br>(82.4 – 100) | 93.7<br>(90.4 – 95.9) | 48.6<br>(33.4 – 64.1) | 100<br>(98.7 – 100) | 94.0<br>(90.9 – 96.2) | 0.63<br>(0.46 – 0.79) |
| qPCRs on minimally processed samples | NP | 4.7 (15)<br>(2.9 – 7.6) | 73.3<br>(48.0 – 89.1) | 91.4<br>(87.8 – 94.1) | 29.7<br>(17.5 – 45.8) | 98.6<br>(96.4 – 99.4) | 90.6<br>(86.9 – 93.3) | 0.38<br>(0.17 – 0.59) |
|  | OP | 15.7 (50)<br>(12.1 – 20.1) | 34.0<br>(22.4 – 47.8) | 92.6<br>(88.8 – 95.1) | 45.9<br>(31.0 – 61.6) | 88.3<br>(84.0 – 91.5) | 83.4<br>(78.9 – 87.1) | 0.30<br>(0.12 – 0.47) |
|  | either NP or OP | 19.1 (61)<br>(15.2 – 23.8) | 41.0<br>(29.5 – 53.5) | 95.3<br>(92.0 – 97.3) | 67.6<br>(51.5 – 80.4) | 87.2<br>(82.8 – 90.6) | 85.0<br>(80.6 – 88.5) | 0.43<br>(0.28 – 0.58) |
| qPCRs on culture-enriched samples | NP | 4.7 (15)<br>(2.9 – 7.6) | 86.7<br>(62.1 – 96.3) | 92.1<br>(88.5 – 94.6) | 35.1<br>(21.8 – 51.2) | 99.3<br>(88.3 – 94.4) | 91.8<br>(88.3 – 94.4) | 0.46<br>(0.27 – 0.66) |
|  | OP | 12.2 (39)<br>(9.1 – 16.3) | 61.5<br>(45.9 – 75.1) | 95.4<br>(92.2 – 97.3) | 64.9<br>(48.8 – 78.2) | 94.7<br>(91.4 – 96.8) | 91.2<br>(87.6 – 93.9) | 0.58<br>(0.43 – 0.73) |
|  | either NP or OP | 16.3 (52)<br>(12.7 – 20.8) | 67.5<br>(53.8 – 78.5) | 99.3<br>(97.3 – 99.8) | 94.6<br>(82.3 – 98.5) | 94.0<br>(90.6 – 96.2) | 94.0<br>(90.9 – 96.2) | 0.75<br>(0.65 – 0.86) |

NP – nasopharyngeal; OP – oropharyngeal; PPV – positive predictive value; NPV – negative predictive value; 95%CI – 95% confidence interval; $\kappa$ – Cohen's kappa where $\leq 0$ , 0.01-0.20, 0.21-0.40, 0.41-0.60, 0.61-0.80, $>0.81$ are interpreted as poor agreement, slight, fair, moderate, substantial, and almost perfect agreement, respectively.

### Comparison of Serotype Carriage Detection Methods

With conventional culture (without PCR-guided additional culture), 29% (445/1549) of nasopharyngeal samples including 45% (422/946) from children and 4% (23/603) from adults were positive for a serotype (nontypeable pneumococci excluded), as already described above. Next, we assessed the accuracy of molecular methods when applied to detect carriage of pneumococcal serotypes. **Figure S1** depicts results of serotype detection in culture-enriched samples tested with serotype-specific qPCR assays. None of the samples generated any signal in qPCRs targeting serotype 1, and 23F and serogroup 18, nor was positive for any of these serotypes by culture. The assays targeting serotypes 4, 5 showed a lack of specificity and the assay targeting serotype 17F showed lack of sensitivity when applied to both, nasopharyngeal and to oropharyngeal samples. Results of these three assays were excluded from analysis.

Four-hundred and twenty-three nasopharyngeal samples were positive for one or more serotypes targeted in qPCRs either by culture (n=393 samples) or with molecular methods (n=411 samples) and applying ^ROCd^C_q_ criterium for positivity for a serotype. It included 42% (395/946) samples from children and 3% (16/603) from adults. Altogether, there were n=479 serotypes carriage events detected by testing nasopharyngeal samples with either conventional culture or molecular methods (**Table S3**). For serotypes targeted by qPCRs detected in nasopharyngeal samples the results of molecular detection displayed excellent agreement (ICC 0.93, 95% CI 0.92-0.94) with *piaB* and *lytA* C_q_s (**Fig. 3**) and, with exception of serotype 3, almost perfect agreement (Cohen’s kappa >0.81) with isolation of live strain of a particular serotype from nasopharyngeal swab (**Fig. 4**). Also, results of serotype detection in oropharyngeal samples from adults displayed excellent agreement with *piaB* and *lytA* C_q_s (ICC 0.96, 95% CI 0.92-0.98) (**Fig. S2**). Finally, there was near-perfect agreement between overall serotypes carriage events detected by qPCR compared with detected by culture (**Table 4**).

**Figure 3.**
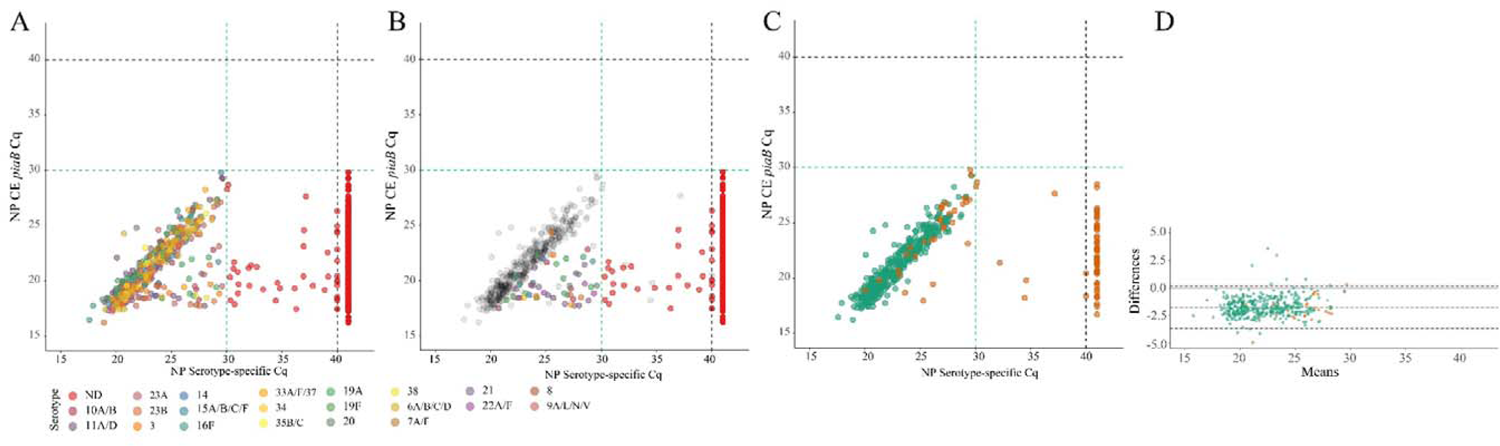
Scatter plots (**A**-**C**) displaying correlation between cycle threshold (C_q_) from real-time PCR (qPCR) assays targeting the *Streptococcus pneumoniae piaB* and serotype/serogroup specific signal detected with qPCR for nasopharyngeal samples classified positive for *S. pneumoniae* according to ^ROCd^C_q_ criterium (green dashed lines). Bland-Altman plot (**D**) displaying agreement between *piaB* and dominant serotype detected by qPCR. Each dot represents an individual serotype carriage event detected by qPCR in culture-enriched nasopharyngeal sample (**A-C**). Dots in panel **A** depicts serotypes detected with qPCR. Dots are color-coded according to serotypes/serogroup targeted in an assay (see legend). Colour dots in panel **B** depict subdominant serotypes detected qPCR while grey dots mark dominant serotypes. Red dots in panel **A** and **B** depict samples not generating any signal in serotype/serogroup-specific qPCRs or with the signal of C_q_ higher than the ^ROCd^C_q_ threshold, hence classified as negative for a serotype. In panels **C** and **D** green dots mark samples with congruent serotype between culture and molecular methods and orange dots mark samples with noncongruent result. In panel **D** shaded grey areas mark the 95% confidence interval of the upper limit of agreement, bias and lower limit of agreement. The solid line grey line marks the line of equality.

**Figure 4.**
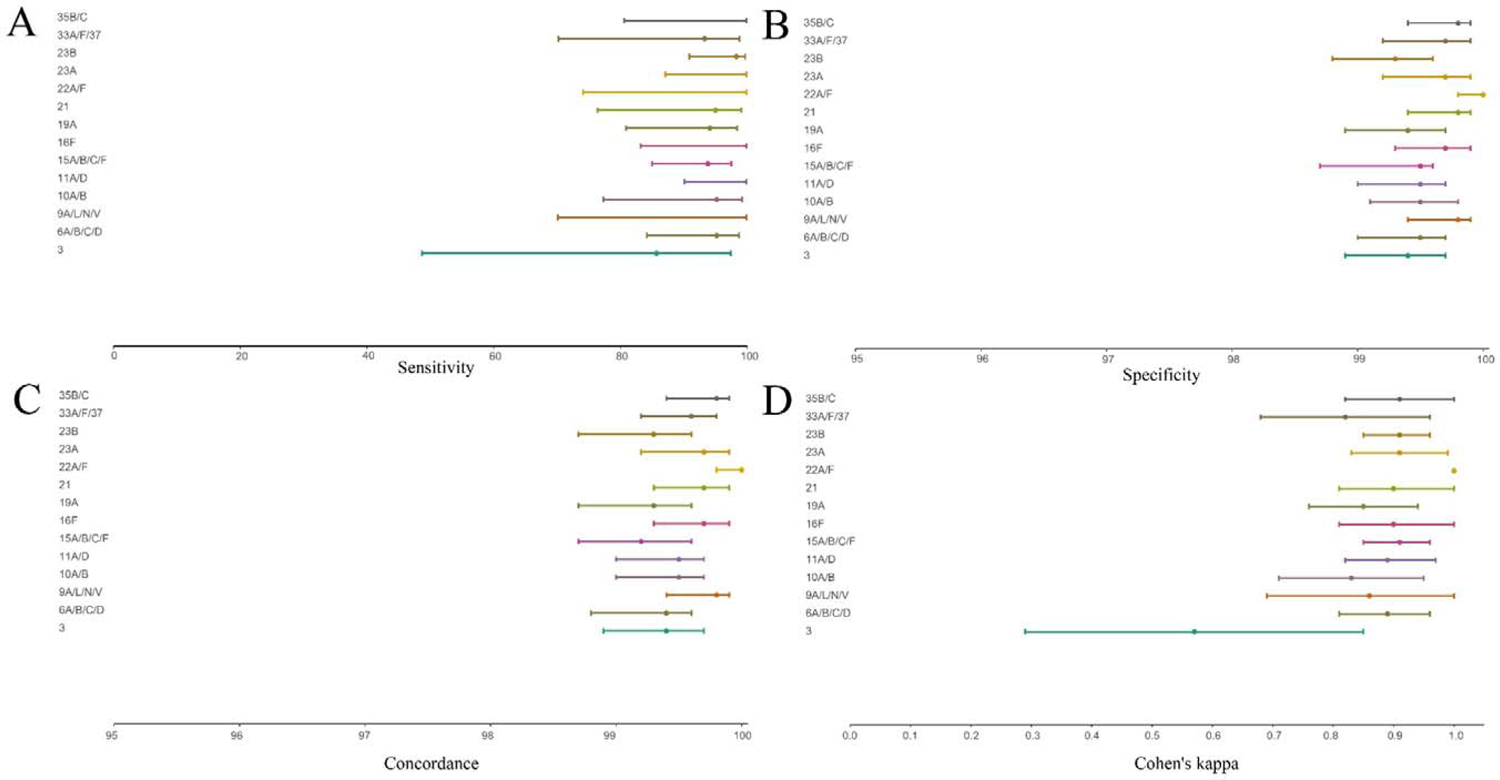
Forest plot displaying the point estimate and 95% confidence of intervals sensitivity (**A**), specificity (**B**), concordance (**C**) and Cohen’s kappa (**D**) for molecular diagnostic tests applied to culture-enriched samples when compared to isolation of *S. pneumoniae* strains of particular serotype/serogroup from nasopharyngeal swabs collected in the study. Graphs displayed results for serotypes/serogroups that have been cultured from >5 nasopharyngeal samples.

**Table 4.** The accuracy of *Streptococcus pneumoniae* **serotype** detection in n=1549 nasopharyngeal samples from children (n=946) and adults (n=603) tested using molecular methods applied to DNA extracted culture-enriched samples. Measures of diagnostic accuracy were calculated by comparing the numbers of serotype carriage events detected with molecular methods with n=393 samples from which serotypes were cultured that were targeted by serotype-specific qPCR assays.

| study population | PPV<br>%<br>(95%CI) | NPV<br>%<br>(95%CI) | Sensitivity<br>%<br>(95%CI) | Specificity<br>%<br>(95%CI) | Concordance<br>%<br>(95%CI) | $\kappa$<br>(95%CI) |
| --- | --- | --- | --- | --- | --- | --- |
| all<br>(n=1549) | 100<br>(99.0 – 100) | 98.7<br>(97.9 – 99.2) | 96.2<br>(93.8 – 97.7) | 100<br>(99.7 – 100) | 99.0<br>(99.7 – 100) | 0.97<br>(0.96 – 0.99) |
| children<br>(n=946) | 98.1<br>(96.6 – 98.9) | 98.1<br>(96.6 – 98.9) | 97.1<br>(94.8 – 98.4) | 100<br>(99.3 – 100) | 98.8<br>(97.9 – 99.3) | 0.98<br>(0.96 – 0.99) |
| adults<br>(n=603) | 100<br>(78.5 – 100) | 99.3<br>(98.3 – 99.7) | 77.8<br>(54.8 – 91.0) | 100<br>(99.3 – 100) | 99.3<br>(98.3 – 99.7) | 0.87<br>(0.75 – 1) |

PPV – positive predictive value; NPV – negative predictive value; 95%CI – 95% confidence interval; $\kappa$ – Cohen’s Kappa where $\leq 0$ , 0.01-0.20, 0.21-0.40, 0.41-0.60, 0.61-0.80, $>0.81$ are interpreted as poor agreement, slight, fair, moderate, substantial, and almost perfect agreement, respectively.

Multiple-serotype carriage events were significantly more often detected using molecular methods compared with culture (*p*<0.0001). Furthermore, despite a limited number of different serotypes tested by qPCR, observed multiple-serotype carriage frequencies were not significantly different from expected frequencies based on molecular detection on culture-enriched NP samples (11% versus 9%, respectively; one proportion Z-test, *p*=0.2669) unlike detection of multiple-serotype carriage by culture which significantly underestimated expected frequencies (2% versus 9%, respectively; one-proportion Z-test, *p*<0.0001).

### Interlaboratory Comparison of Methods

To evaluate the reproducibility of molecular methods and to assess the agreement in laboratory results between the Netherlands and England we processed a subset of culture-enriched samples from both countries in both laboratories. Results for *piaB* and *lytA* qPCRs demonstrated good reliability between both laboratories (**Table S4**). We observed near-perfect agreement identifying culture-enriched samples as positive for pneumococcus with molecular methods, and substantial agreement for minimally processed nasopharyngeal samples (**Table S5**). For culture-enriched nasopharyngeal samples we evaluated agreement between both laboratories for serotype carriage detection by qPCR. Overall, near-perfect agreement was observed (Cohen’s kappa 0.82, 95% CI 0.74 – 0.90).

## DISCUSSION

In the current study we have demonstrated that molecular methods exhibit near-perfect agreement to conventional culture in the detection and serotyping of *S. pneumoniae* in children and adults when a nasopharyngeal swab is the only sample tested. Furthermore, we have observed increased sensitivity of *S. pneumoniae* carriage detection among adults by testing oropharyngeal samples with molecular methods and conducting qPCR-guided culturing. We highlight several statistical procedures that can be used to evaluate the reliability of molecular results and enhance the specificity of molecular methods for the detection of live *S. pneumoniae*.

The current gold standard method for carriage detection is the isolation of live pneumococci from cultures of deep trans-nasal nasopharyngeal swab, in adults complemented with culture of a swab collected trans-orally (OP) [4]. However, the gold standard lacks sensitivity in case of low-density carriage or when applied to poly-microbial samples in which *S. pneumoniae* is not a dominant bacterium and it does not allow the detection of co-carriage of pneumococcal strains [7, 18, 19, 21, 43]. Since the density of pneumococcal carriage episodes in adults is much lower than in children [18], sampling multiple sites increases sensitivity of carriage detection [7, 16, 18]. However, carriage surveillance based exclusively on primary diagnostic cultures of nasopharyngeal and oropharyngeal samples often provides low quality data in adults. This limitation of the gold standard method is of particular concern for surveillance of carriage in older adults [19, 24], the age group with the largest incidence and burden of pneumococcal pneumonia and invasive pneumococcal disease [2, 44] as carriage is often reported to be virtually absent when the gold standard method is applied [16, 45].

To overcome these limitations, we complement conventional culture with molecular methods to improve the overall sensitivity of *S. pneumoniae* carriage detection [7, 18, 21–24]. This approach is particularly effective when applied to highly poly-microbial samples from the oral niche. To validate this method, we have compared the performance of molecular methods to the gold standard. While demonstrating near-perfect agreement with primary nasopharyngeal cultures from children, application of qPCR-based methods still significantly increased the number of carriers detected. Near perfect agreement was also observed in adults when analysis focused explicitly on nasopharyngeal swabs. In adults, application of qPCR-based methods to nasopharyngeal samples did not increase sensitivity of carriage detection.

Importantly, in adults testing oropharyngeal samples changed the results dramatically. By revisiting samples negative in primary culture yet positive by qPCR with culture qPCR-guided culturing we have significantly increased the number of adult carriers from whom viable pneumococci were isolated. It demonstrates that molecular methods can improve the overall sensitivity of *S. pneumoniae* detection, a result in line with previous reports by us and others [7, 16, 18, 22, 46]. It also highlights that the gold standard method applied to oropharyngeal cultures severely underestimates presence of *S. pneumoniae* in adults as qPCR-guided culturing increased by 7.7-fold the number of oropharyngeal samples from which viable pneumococci were isolated. Interestingly, there is evidence that testing oropharyngeal in addition to nasopharyngeal samples substantially enhances sensitivity of carriage detection also in children [17].

Concerns have been raised that the molecular methods are overly sensitive and lack specificity for the detection of live bacteria as “relic DNA” (DNA from non-intact cells) could be detected as well [28]. Indeed, live pathogens are less likely to be cultured from samples displaying weak positivity by qPCR [21, 47]. While this may reflect limitations in current culturing techniques, it could also indicate presence of relic DNA that may reflect recent exposure to pneumococcus instead of colonization proper [28]. The addition of the culture-enrichment procedure prior to molecular detection reduces the risk of misclassifying these events as carriage, thus improving the specificity of detection for live pneumococci. Culture-enrichment also increases the sensitivity of carriage surveillance, in particular for poly-microbial samples [7, 18, 21–24, 46–48]. Indeed, we observed that *S. pneumoniae* detection in culture-enriched samples displayed significantly increased, sensitivity, specificity, and agreement with culture and qPCR-guided culture when compared with detection in minimally processed samples.

To further improve the specificity of molecular methods for live pneumococci we performed ROC curve analysis to estimate with the Youden index C_q_ cut-off values that display an optimal combination of sensitivity and specificity for samples positive for *S. pneumoniae* by culture in primary cultures or qPCR-guided cultures [37]. This approach excluded samples that exhibited minimal positivity for targeted genes by qPCR or displayed poor agreement between *piaB* and *lytA* genes as shown in Bland-Altman graphs and with the intra-class correlation coefficient. The use of the Youden index in a ROC curve analysis to identify optimal C_q_ cut-off values enhances the specificity of molecular methods for detection of live pneumococcus, and the ^ROCd^C_q_ criteria can be used to direct qPCR-guided culturing. The success of this approach is dependent on culturing methods, therefore care should be taken to employ sensitive culturing techniques, such as the use selective culture plates [4]. Furthermore, for molecular methods to be informative for culturing efforts DNA extraction should be performed irrespective of the identification of pneumococcal growth in cultures. ROC curve analysis with the Youden index may yield overly stringent C_q_ cut-off values if qPCR-guided culturing efforts are not conducted, which could result in classifying samples containing live pneumococci as negative by qPCR. Recent advances in culturing techniques may further enhance the specificity of molecular methods for the detection of live *S. pneumoniae* [49].

Some studies have cautioned against the use of molecular methods due to the presence of pneumococcal genes among commensal streptococci [25–27]. This phenomenon is likely to be common among bacterial species co-existing in a shared niche [50–52] and of particular concern for highly poly-microbial samples, hence careful selection of targeted genes is important [53]. As previously described by our group [18, 22, 23] and others [54] a “two-to-tango” approach, quantifying both *piaB* and *lytA* genes with molecular methods enhances the specificity of *S. pneumoniae* detection in poly-microbial samples. This approach allows for measurements to be evaluated in a reliability analysis on a per sample basis [39] and for bias between measurements to be identified. While systemic biases between quantified genes could reflect dissimilarities between qPCR assay efficiencies, non-systemic biases can arise due to the presence of a gene or closely related sequence in DNA from other bacterial species. In case of multiple serotype carriage, serotype-specific abundance of the dominant serotype should display high agreement to targeted genes used to detect pneumococcus (e.g., *piaB* and *lytA*) while serotype-specific abundances of nondominant serotypes may display reduced agreement. Serotypes that exhibit greater abundances than *piaB* or *lytA*, or appear to be present in samples classified as negative for *S. pneumoniae* are likely to be due to commensal streptococci harbouring genes involved in the biosynthesis of the pneumococcal polysaccharide capsule [55, 56]. As such, certain serotype-specific assays may be nonreliable in poly-microbial sample types.

Insights into the co-occurrence of multiple serotypes in carriage is critical for understanding the dynamics of the serotypes during colonization, host-to-host transmission, and carriage progression into disease. Detection of secondary strains present in co-carriage is also important when distinguishing between unmasking and serotype replacement in assessment of pneumococcal vaccines impact [57]. However, as demonstrated in our study, and described previously the gold standard method does not readily allow detection of multiple serotypes, an event that is likely to occur often in carriage [57]. Furthermore, using our methodology we have observed no significant difference in observed and expected frequencies of multiple-serotype carriage despite what has been reported previously by others whom exclusively used data from conventional culture as method of detection [58].

An important strength of our study is the application of the procedure to two different carriage studies conducted in two different countries, and the evaluation of reproducibility in an interlaboratory comparison. Another strength of our study is that the procedure described is flexible and can be readily adapted to the carriage surveillance of other bacteria, such as *Neisseria meningitidis* [47].

Our study had a number of limitations, the impact of testing oropharyngeal samples was only evaluated for Dutch adults and not adults sampled in England nor for any children. Since oropharyngeal samples tested were collected explicitly from Dutch adults, our findings could be unique for that demographic group and geographic location. In the protocol described we only used *piaB* and *lytA* to detect *S. pneumoniae* with molecular methods and we did not evaluate the procedure with alternative targets described by others [54, 59]. Furthermore, we tested for a limited number of serotypes by qPCR. Finally, not all serotypes were shown to be equally reliably detected with molecular methods and for a number of qPCR assays we were not able to identify the serotype within a serogroup.

In summary, we argue that accurate detection of pneumococcal carriage using qPCR requires concordant quantification of two genes (‘two-targets-to-tango’) to classify a sample as positive for pneumococcus. Similarly, qPCR-based detection requires concordance between pneumococcal and serotype-specific quantification to assure specificity of the method (two-targets-to-tango). We provide evidence that accurate detection of pneumococcal carriage in adults requires at least testing of both, nasopharyngeal and oropharyngeal samples (‘two-samples-to-tango’) and requires molecular detection to be intertwined with culture (‘two methods-to-tango’). Finally, we advise revisiting samples for qPCR-guided culturing (‘two-cultures-to-tango’) when positive by qPCR but negative at primary diagnostic culture. The use of qPCR-guided culturing is of utmost importance for oropharyngeal swabs.

We have outlined the procedure that enhances the specificity of molecular methods for the surveillance of pneumococci and of pneumococcal serotypes in nasopharyngeal and oropharyngeal samples. Our results demonstrate near-perfect agreement between conventional culture and molecular methods when applied to nasopharyngeal samples from children. We have shown that the sensitivity of *S. pneumoniae* carriage surveillance can be greatly enhanced by complementing conventional culture with qPCRs. In adults, testing oropharyngeal on the top of nasopharyngeal samples was of paramount importance for accuracy of pneumococcal carriage detection. Studies investigating impact of testing oropharyngeal samples on detection of pneumococcal carriage in children are needed.

## Authors’ contribution

E.A.M.S. and K.T. had an idea and initiated the study. N.R., E.M., N.K.F., E.A.M.S. and K.T. secured financial support. N.F. and K.T. led the project. A.J.W-M., P.B., C.S., M.v.H., N.R., E.M., and N.K.F. conducted carriage studies, collected the data and provided study materials. W.R.M., J.v.V., and K.T. developed and validated laboratory methods, and wrote the laboratory protocol. W.R.M., J.v.V., D.L., P.B., T.N., S.M., R.T., S.E., and C.S. analysed samples and collected the data. W.R.M., R.M., and K.T. contributed analytical tools. W.R.M., J.v.V., and D.L. curated the data. W.R.M., D.L., N.K.F., and K.T. managed the study. W.R.M., and K.T. performed formal analysis of study data, visualized presentation of the results and drafted the manuscript. All authors amended, critically reviewed, and commented on the final manuscript.

## Data Availability

All data produced in the present work are contained in the manuscript.

---

**Supplementary Figure S1.**
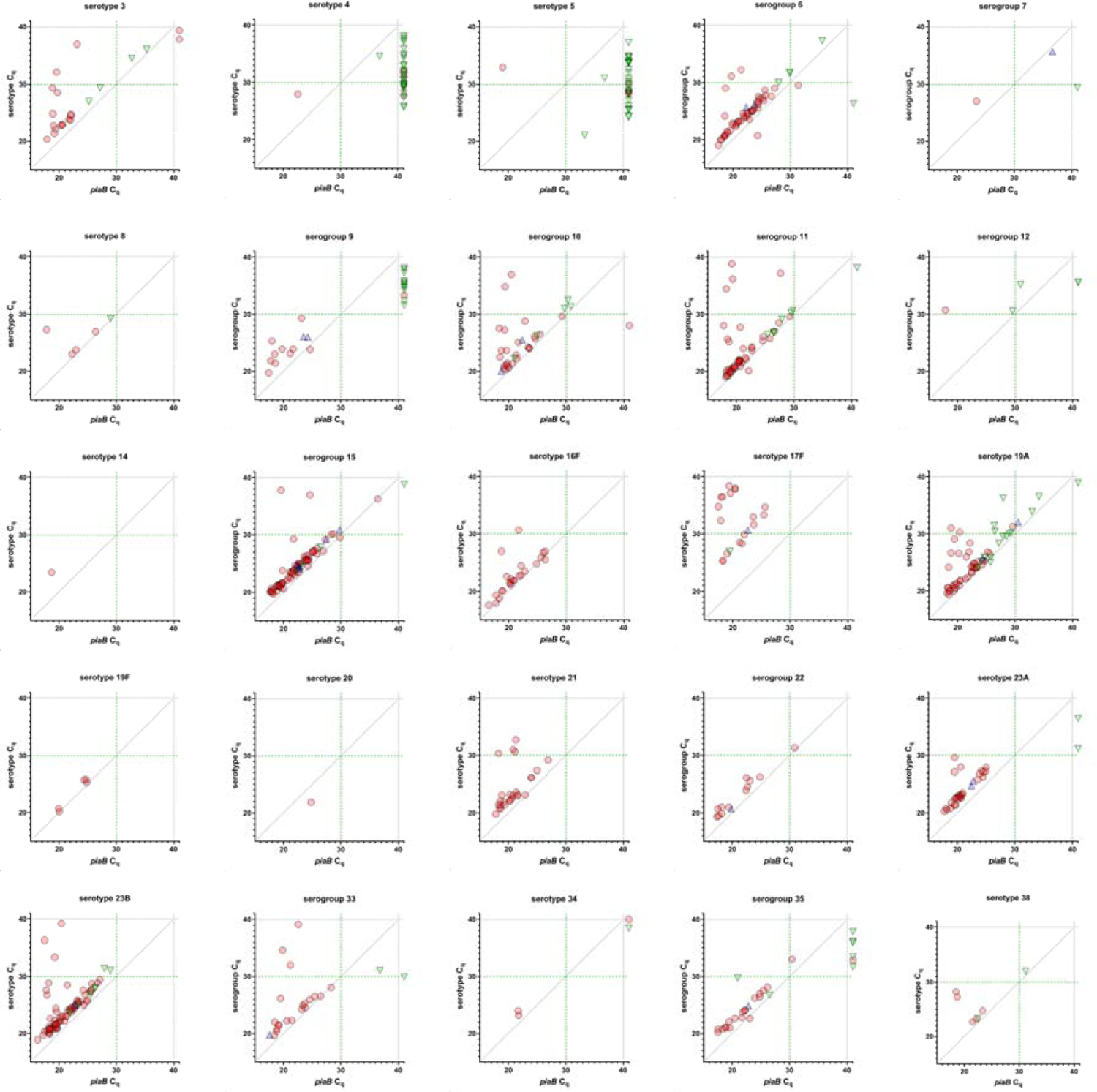
Results of molecular method based quantification of serogroups/serotypes-specific sequences in DNA from culture-enriched nasopharyngeal and oropharyngeal samples. Individual scatter plots depict results of a single serotype-specific or serogroup-specific qPCR assay as labelled above the panel. Each symbol represents an individual sample: pale-red dots represent nasopharyngeal samples from children; blue triangles and green triangles represent nasopharyngeal and oropharyngeal samples from adults, respectively. Sample was classified as positive for a serotype/serogroup with molecular method when the signal detected by qPCR for *piaB* (X-axis) and serotype/serogroup (Y-axis) were both below the ^ROCd^C*_q_* cut-off threshold of 30 C*_q_*. Only samples that generated a signal of C*_q_*<40 in a particular serogroup/serotype-specific qPCR are depicted. Symbols of C*_q_* >40 for *piaB* depict individual samples or pools of samples negative for pneumococcus by qPCR. None of the samples generated any signal in qPCRs targeting serotype 1, serotype 23F and serogroup 18. In addition, no samples were identified as positive for serogroup 12 according to study criteria.

**Supplementary Figure S2.**
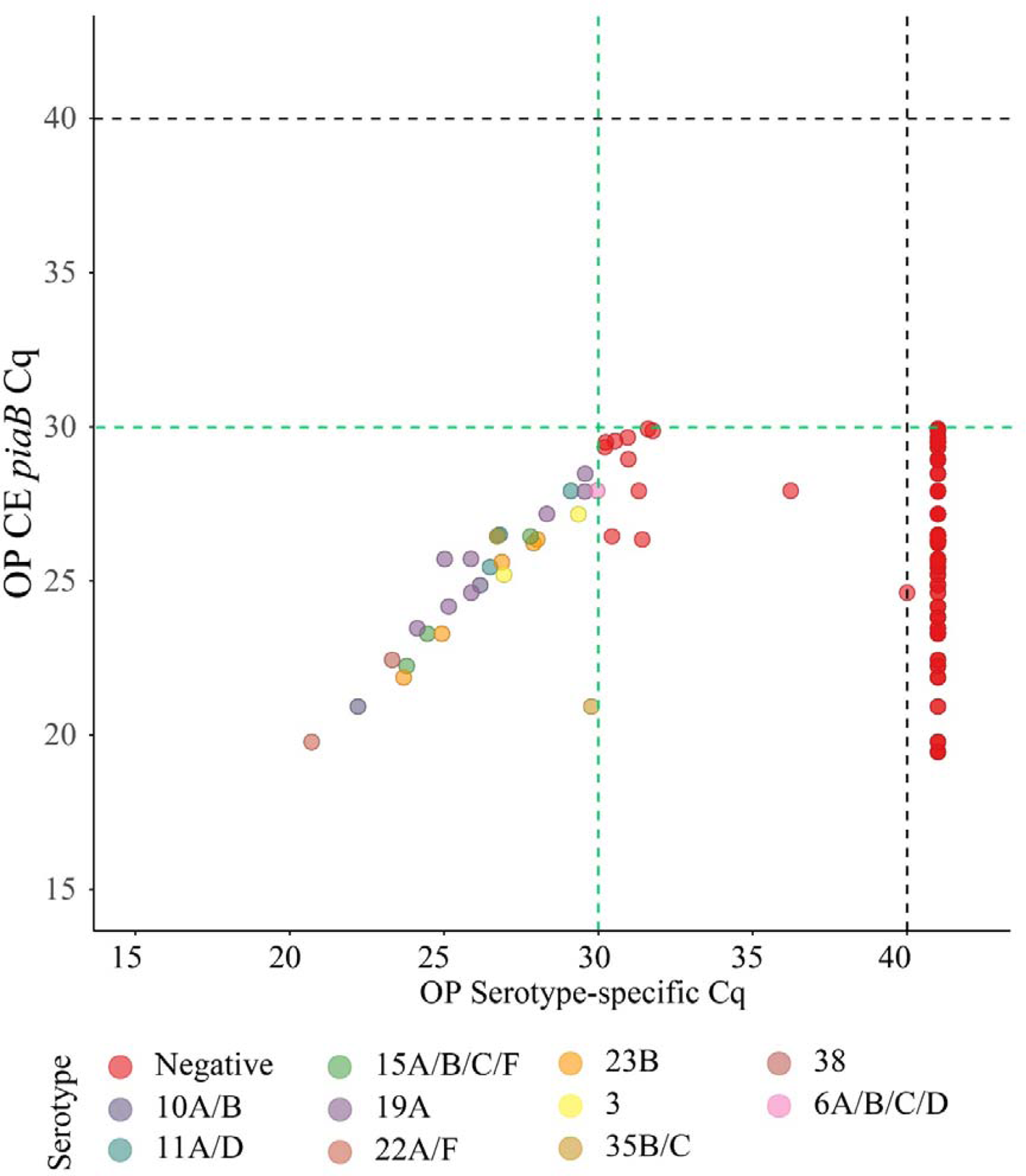
Scatter plot displaying correlation between cycle threshold (C_q_) from real-time PCR (qPCR) assays targeting the *Streptococcus pneumoniae piaB* and serotype/serogroup specific signal detected with qPCR for culture-enriched (CE) oropharyngeal samples from adults classified positive for *S. pneumoniae* according to ^ROCd^C_q_ criterium (green dashed lines). Dots depicts serotypes detected with qPCR. Dots are color-coded according to serotypes/serogroup targeted in an assay (see legend).

**Supplementary Table S1.** Methodological differences between *Streptococcus pneumoniae* carriage studies conducted in the Netherlands and in England.

| Processing step | The Netherlands | England |
| --- | --- | --- |
| transport medium | 1 ml Amies | 1 ml STGG |
| transport temperature | ambient | 2-8°C |
| duration of transport | <8 hours | <48 hours |
| arrival to the diagnostic lab | culturing on receipt | storage at -80°C |
| culture medium | BA-GENT and CBA | COBA and CBA |
| inoculum volume | 35 ul | 50 ul |
| identification<br>of *S. pneumoniae* | optochin-susceptibility & bile solubility tests | bioinformatics-based method applied to bacterial DNA |
| minimally processed (MP)<br>sample | an aliquot of Amies stored at -80°C | an aliquot of skim milk tryptone glucose and glycerine broth medium (STGG) stored at -80°C |
| culture-enriched (CE) sample | whole BA-GENT plate colony growth washed with 2.1 ml of BHI with 10% glycerol | swab of α-haemolytic colonies from COBA (first choice) or CBA, and into 1 ml of STGG |

STGG: skim milk tryptone glucose and glycerine broth medium, CBA: Columbia blood agar plate, BA-GENT: blood agar plate gentamycin, COBA: blood agar plate with *Streptococcus* selective medium.

**Supplementary Table S2.** Results of an intraclass correlation coefficient (ICC) reliability analysis for ^ROCd^C_q_-based criteria in a subset of samples classified as positive for *S. pneumoniae* according to the ^A^C_q_ criterium of <40.

| Sample | $C_q$ score | Number of samples | ICC<br>(95% CI) | p-value |
| --- | --- | --- | --- | --- |
| Minimally processed nasopharyngeal | $< ^{ROCd}C_q$ | 512 | 0.96<br>(0.94-0.97) | < 0.0001 |
| | $> ^{ROCd}C_q$ | 49 | -0.15<br>(-0.42-0.14) | 0.845 |
| Culture-enriched nasopharyngeal | $< ^{ROCd}C_q$ | 469 | 0.92<br>(0.73-0.97) | < 0.0001 |
| | $> ^{ROCd}C_q$ | 10 | -0.19<br>(-0.59-0.41) | 0.751 |
| Minimally processed oropharyngeal | $< ^{ROCd}C_q$ | 59 | 0.96<br>(0.93-0.97) | < 0.0001 |
| | $> ^{ROCd}C_q$ | 24 | -0.323<br>(-0.62-0.08) | 0.947 |
| Culture-enriched oropharyngeal | $< ^{ROCd}C_q$ | 39 | 0.99<br>(0.99-1) | < 0.0001 |
| | $> ^{ROCd}C_q$ | 19 | 0.31<br>(-0.19-0.66) | 0.104 |

ICC: intraclass correlation coefficient - a single score intraclass correlation coefficient with two-way model was used for reliability analysis. ICC values <0.50, 0.50-0.75, 0.75-0.90 and >0.90 are indicative of poor, moderate, good, and excellent reliability, respectively.

**Supplementary Table S3.**
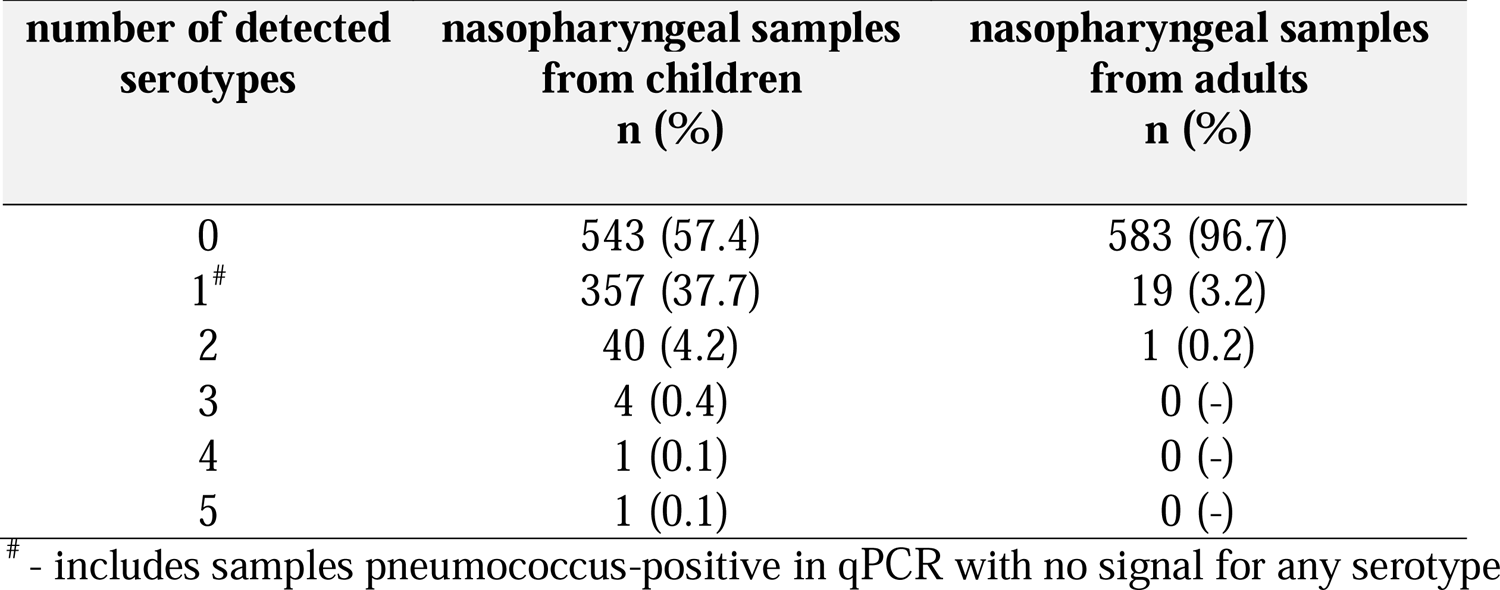
Number of serotypes/serogroups detected with any method.

| **number of detected**<br>**serotypes** | **nasopharyngeal samples**<br>**from children**<br>**n (%)** | **nasopharyngeal samples**<br>**from adults**<br>**n (%)** |
| --- | --- | --- |
| 0 | 543 (57.4) | 583 (96.7) |
| 1<sup>#</sup> | 357 (37.7) | 19 (3.2) |
| 2 | 40 (4.2) | 1 (0.2) |
| 3 | 4 (0.4) | 0 (-) |
| 4 | 1 (0.1) | 0 (-) |
| 5 | 1 (0.1) | 0 (-) |

<sup>#</sup> - includes samples pneumococcus-positive in qPCR with no signal for any serotype

**Table S4.**
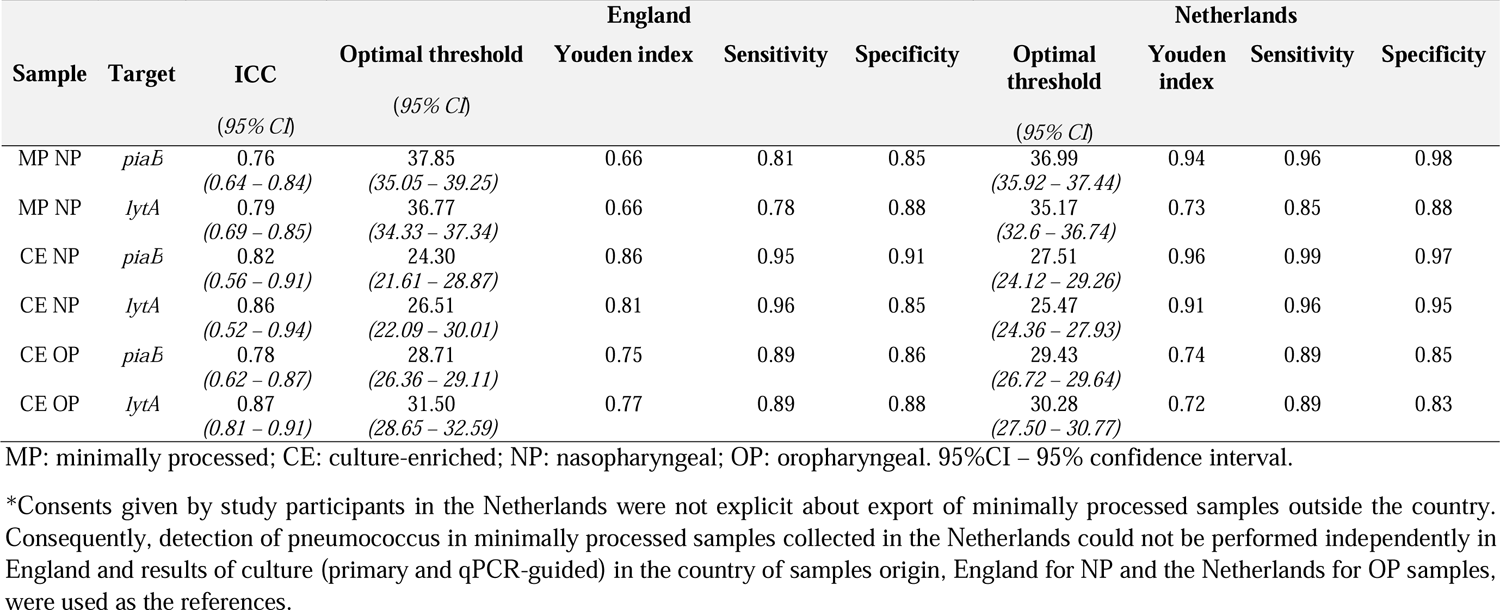
Optimal qPCR cycle threshold C_q_ and corresponding parameters for *Streptococcus pneumoniae* carriage detection in a subset of nasopharyngeal (n=176) and oropharyngeal (n=100) samples processed independently in England and in the Netherlands. Results from qPCR were validated in a receiver operating characteristic curve analysis with culture as reference*.

**Table S5.** The accuracy of *Streptococcus pneumoniae* detection in n=176 nasopharyngeal samples and n=59 oropharyngeal tested independently in both countries using molecular methods applied to DNA extracted from minimally processed and culture-enriched samples and applying ^ROCd^C_q_ thresholds for a sample positivity in qPCRs. Measures of diagnostic accuracy were calculated by comparing the number of detected nasopharyngeal and oropharyngeal samples positive per method with the number of individuals positive for *S. pneumoniae* based on isolation of live pneumococcus either from the primary diagnostic or qPCR-guided culture (n=101 of nasopharyngeal samples culture-positive in UK and n=19 oropharyngeal samples culture-positive in the Netherlands).

| Method | Percent (n) of samples tested positive in the Netherlands<br>(95%CI) | Percent (n) of samples tested positive in England<br>(95%CI) | Concordance %<br>(95%CI) | $\kappa$<br>(95%CI) |
| --- | --- | --- | --- | --- |
| qPCRs on minimally processed NP samples | 42.0 (74)<br>(35.0 – 49.4) | 54.5 (96)<br>(47.2 – 61.7) | 83.0<br>(76.7 – 87.8) | 0.66<br>(0.55 – 0.77) |
| qPCRs on culture-enriched NP samples | 59.1 (104)<br>(51.7 – 66.1) | 55.1 (97)<br>(47.7 – 62.3) | 93.8<br>(89.2 – 96.5) | 0.87<br>(0.80 – 0.95) |
| qPCRs on culture-enriched OP samples | 22.0 (13)<br>(13.4 – 34.1) | 25.4 (15)<br>(16.1 – 37.8) | 98.0<br>(93.0 – 99.4) | 0.95<br>(0.88 – 1) |

95%CI – 95% confidence interval; $\kappa$ – Cohen’s Kappa where $\leq 0$ , 0.01-0.20, 0.21-0.40, 0.41-0.60, 0.61-0.80, $>0.81$ are interpreted as no agreement, none to slight, fair, moderate, substantial, and almost perfect agreement, respectively.

